# Persistent and reversible impacts of smoking on resting-state EEG in chronic smokers and successful long-term abstainers

**DOI:** 10.1101/2022.06.19.22276601

**Authors:** Hyeji Lee, Yoonji Jeon, Cheolin Yoo, HeeYoung Seon, Jiwon Park, Minho Hwang, Kwangyeol Baek, Dongil Chung

**Affiliations:** Department of Biomedical Engineering, Ulsan National Institute of Science and Technology, Ulsan, South Korea; Department of Biomedical Convergence Engineering, Pusan National University, Busan, South Korea; Department of Occupational and Environmental Medicine, Ulsan University Hospital, University of Ulsan College of Medicine, Ulsan, South Korea; Department of Psychology, The University of Edinburgh, Edinburgh, United Kingdom

## Abstract

Smoking is a severe addictive health risk behavior and notorious for the high likelihood of relapse after attempted cessation. Such an addictive pattern in smoking has been associated with neurobiological changes in the brain. However, little is known whether the neural changes associated with chronic smoking persist after a long period of successful abstinence. To address this question, we examined resting state EEG (rsEEG) in heavy smokers who have been smoking for 20 years or more, past-smokers who have been successfully abstaining for 20 years or more, and non-smokers. Compared with chronic current- or past-smokers, non-smokers showed higher relative power in theta frequency band, showcasing long-lasting effects of smoking on the brain. A few rsEEG features in alpha frequency band also revealed reversible impacts of smoking, such that only current-smokers, but not past-smokers, showed distinctively higher patterns than non-smokers in their relative power, EEG reactivity—power changes between eyes-closed and eyes-open conditions—, and coherence between channels. Furthermore, rsEEG feature differences between current- and past-smokers were accounted for by individuals’ self-reported smoking history and nicotine dependence. These data suggest long-lasting impacts of chronic smoking on the brain that are dissociable from the neural changes reversible with long-term abstinence.

## Introduction

Smoking is one of the most severe addictive health risk behaviors and the leading cause of preventable death that accounts for more than 8 million deaths [1]. A distinctive characteristic observed in smokers is that a large proportion of individuals who attempt to quit, particularly the ones who already attempted before [2], is highly likely to relapse [3]. Such an addictive pattern has been described as a “cycle of spiraling dysregulation” of brain [4] suggesting that neurobiological changes in the brain [5-7] account for the reason why individuals become addicted and why they are vulnerable to relapse. This perspective was corroborated by previous studies showing that among many factors, pronounced cue-induced neural responses [8-11] and smaller brain volume observed in smokers are associated with a high risk of relapse [12]. A natural question that follows is whether or not smoking has long-term and long-lasting effects on the brain. However, it still remains elusive whether these changes in the brain of smokers are permanent even after a long period of successful abstinence, or reversible at least to some extent. To address this issue, both the impacts of smoking and of abstinence need to be examined from individuals who have different histories of lifetime smoking.

Previous studies showed that electroencephalography (EEG) can be used to capture neural characteristics in psychopathology [13,14]. Among various features in EEG, the differences in spontaneous and intrinsic brain activities measured under the resting state [15] have been found useful in dissociating smokers from non-smokers [16-18], and in characterizing smokers’ abstinence [19-21] and craving severity [22]. Particularly, being a smoker or administrating nicotine has been typically associated with decreased EEG powers in delta or theta frequency bands [16,17], but with increased powers in alpha band [23-25]. Note that most studies focused on the impact of smoking (acute or chronic) or a brief nicotine deprivation. The aim of the current study is to examine neural changes in individuals who have different smoking profiles and dissociate reversible from long-lasting (or potentially irreversible) impacts. Based on previous reports on EEG biomarkers associated with smoking, we hypothesized that resting state EEG (rsEEG) powers measured from smokers, past-smokers, and non-smokers would reveal irreversible neural changes associated with chronic smoking apart from reversible changes with successful long-term abstinence.

To examine long-term effects of smoking and abstinence, we explored EEG coherence as an additional feature in EEG. Previous studies showed that individuals with substance dependence (e.g., nicotine [26], cannabis [27], alcohol [28], and heroine [29]) or behavioral addiction (e.g., gaming disorder and internet addiction [30]) demonstrate altered EEG coherence. In a recent study, Prashad et al. [27] reported EEG coherence patterns in cannabis users, and suggested a possibility of using EEG coherence as a biomarker for substance use disorders. Winterer et al. [28] examined EEG coherence in long-term abstinent alcohol users and showed that an increase in EEG coherence may be a trait-like (i.e., irreversible or invariant) feature for substance abuse. Based on these previous reports, we expected EEG coherence to provide a useful measure for investigating both the long-term effects of smoking and abstinence.

In a series of studies examining the impact of neurodegenerative changes in the brain (e.g., Alzheimer’s disease), alpha reactivity—relative reduction in alpha band EEG power during eyes-open compared to eyes-closed resting state—has been suggested as a biomarker of cholinergic system integrity [31-33]. Specifically, a recent study suggested that individuals’ alpha reactivity levels are associated with their functional connectivity between the visual cortex and the nucleus basalis of Meynert (NBM) [32], a brain region considered as the main source of cortical cholinergic innervation. Given that chronic nicotine exposure is known to induce increases in the number of nicotinic acetylcholine receptors (nAChR) and losses in their functional sensitivity [34,35], we expected that chronic cigarette smoking may affect functional connectivity between the associated brain regions and EEG patterns which are suggested to reflect the relationship. Across various previous studies that examined the relationship between smoking and alpha rhythm EEG, the focus has been on the impact of acute nicotine administration or temporary deprivation [19,20,24,36] and of cue-induced responses [10,37]. Here, based on previously reported neurobiological changes linked to chronic smoking, we hypothesized that alpha reactivity in addition to EEG power and coherence may reflect long-term effects of smoking.

Data from 20 chronic heavy smokers who at least had been smoking a pack of cigarettes for 20 years, 28 past-smokers who successfully stayed abstinent for 20 years, and 33 non-smoker controls (non-smoker hereafter) were analyzed for the current study. To characterize and investigate the impacts of cigarette smoking on the brain, participants’ rsEEGs recorded their eyes-open and closed were analyzed. If there exist irreversible neural alterations associated with smoking, similar neural patterns should be observed between smokers and past-smokers even if they have been successfully staying abstinent for a long period, whereas the patterns should be different from that of non-smokers. To further examine individual differences, we explored correlations between individuals’ EEG features (power, coherence, and alpha reactivity) and their smoking related self-report measures (e.g., cigarettes per day, nicotine dependence, and the number of quit attempts).

## Material and Methods

### Participants

Twenty-five chronic smokers, 38 non-smokers, and 31 past-smokers were recruited for the current study. Smokers were recruited from a one-week group behavior therapy for smoking cessation, organized at Ulsan University Hospital, and all the smokers have been smoking at least one pack of cigarettes per day for 20 years (20 pack-years). The exclusion criteria included neurological or psychiatric disorder, traumatic brain injury, and current use of psychoactive substances other than nicotine, and were initially assessed at the sign-up interview for the current study. Only three out of 25 smokers who met the inclusion criteria were female, which reflects higher prevalence of male than female smokers in South Korea [38]. Due to this gender bias, we decided to exclude these female participants from the analyses and left it as a limitation of the current study (see **Discussion**). Two additional smokers were excluded due to large EEG artifacts (see **EEG analysis** for artifact definition). At study intake, all smoker participants were abstinent for at least 24 hours and only five participants smoked 6.20 ± 5.63 cigarettes in last 48 hours. Before the EEG measurement, participants’ abstinence was confirmed by the exhaled carbon monoxide (eCO) level of 3 ppm or below (Micro Smokerlyzer, Bedfont Scientific Ltd., Maidstone, Kent, UK; BMC-2000, Senko International Inc., Osan-si, Gyeonggi-do, Republic of Korea). Non-smokers and past-smokers were recruited from local community via online advertisements. Non-smokers were defined as individuals who smoked < 5 cigarettes in their lifetime and had not smoked in last month. To examine the effect of chronic smoking that persist after a long period of abstinence, we set inclusion criteria for the past-smoker group as following: 1) regularly smoked 10 or more cigarettes per day at least for 6 months, and 2) has been ceased smoking for 20 years or more. As in the smoker group, none of past- or non-smokers met the following exclusion criteria: history of neurological disorder, psychiatric disorder, or traumatic brain injury, and current use of psychoactive substances. Past- and non-smokers were also confirmed that they showed eCO level of either 0 or 1 ppm. Three past-smokers and five non-smokers were excluded due to extreme EEG artifacts (see **EEG analysis** for artifact definition). After applying aforementioned exclusion/inclusion criteria, final sample included 20 smokers (age = 56.30 ± 6.93), 28 past-smokers (age = 54.64 ± 6.36), and 33 non-smokers (age = 53.09 ± 4.88). See **Table 1** for additional demographic information and smoking histories. The current study and all the protocols were approved by the Institutional Review Board (IRB) of Ulsan National Institute of Science and Technology (UNIST) (UNISTIRB-19-45-A), and all participants provided written informed consent following an explanation of study procedures.

**Table 1.** Demographic information and smoking measures.

|  | Non-smoker<br>(n = 33) | Past-smoker<br>(n = 28) | Smoker<br>(n = 20) | Test statistics |
| --- | --- | --- | --- | --- |
| Age (years) | 53.09 ± 4.96 | 54.64 ± 6.48 | 56.30 ± 7.12 | F(2, 78) = 1.76<br>P = 0.18 |
| Education<sup>a</sup> | 3.67 ± 1.14 | 3.32 ± 1.12 | 3.00 ± 0.97 | $\chi^2(8) = 14.37$<br>P = 0.073 |
| Income level<sup>b</sup> | 3.15 ± 1.25 | 3.14 ± 1.41 | 2.85 ± 1.42 | F(2, 78) = 0.37<br>P = 0.69 |
| K-BDI-II | 8.63 ± 6.89 | 9.96 ± 6.50 | 8.75 ± 6.27 | F(2, 78) = 0.35<br>P = 0.71 |
| Age of smoking initiation** | - | 19.43 ± 1.32 | 18.10 ± 1.80 | t(46) = -2.95<br>P = 0.0049 |
| Years smoking*** | - | 10.21 ± 6.43 | 37.80 ± 7.54 | t(46) = 13.63<br>P = 9.07e-18 |
| Cigarettes per day<sup>c</sup> | - | 18.24 ± 9.05 | 19.50 ± 8.02 | t(39) = 0.47<br>P = 0.64 |
| Quit attempts<sup>c</sup> | - | 2.11 ± 2.11 | 2.15 ± 1.04 | U = 142.00<br>P = 0.12 |
| Abstinence years | - | 24.84 ± 4.70 | - | - |
| FTND total**<sup>c</sup> | - | 3.15 ± 3.17 | 5.58 ± 2.09 | t(37) = 2.81<br>P = 0.0079 |
| QSU-brief***<sup>c</sup> | - | 10.80 ± 2.73 | 21.32 ± 10.17 | t(37) = 4.46<br>P = 7.30e-05 |

Means ± SDs are reported for each item. K-BDI-II, Korean Version of Beck Depression Inventory, Second Edition [43]; FTND, Fagerström Test for Nicotine Dependence [39,40]; QSU-brief, brief Questionnaire of Smoking Urges [41]; <sup>a</sup>Average education where 1 = ≤ middle school graduate, 2 = high school graduate, 3 = some university or community college graduate, 4 = Bachelor's degree, 5 = ≥ Postgraduate studies; <sup>b</sup>Average household monthly income where 1 = ≤ \$1800, 2 = \$1800 to 3500, 3 = \$3500 to 5300, 4 = \$5300 to 7000, 5 = \$7000 to 8800, and 6 = ≥ \$8800; <sup>c</sup>Quit attempts data are missing from ten past-smokers, and cigarettes per day data are missing from seven past-smokers. FTND and QSU-brief data are missing from one smoker and eight past-smokers; \*\*P < 0.01, \*\*\*P < 0.001.

### Smoking history and clinical measures

Smokers and past-smokers completed a battery of self-report measures, assessing their smoking history, nicotine dependence, and craving for nicotine. Nicotine dependence was measured using the Fagerström Test for Nicotine Dependence (FTND) [39,40], and craving for nicotine was measured using a brief, 10-item version of the Questionnaire of Smoking Urges (QSU-brief) [41]. Given a tight association between smoking cessation attempts and depression [42], individuals’ depression symptoms were also measured using Korean version of Beck Depression Inventory, second edition (K-BDI-II) [43].

### EEG acquisition

The electroencephalogram (EEG) was digitized at a sampling frequency of 500 Hz, amplified using the actiCHamp system (Brain Products GmbH, Germany), and recorded from 31 Ag/AgCl electrodes mounted in a cap (actiCap, Brain Products GmbH, Germany). We used the Fz electrode as a reference and the FPz as a ground. Impedances of all electrodes were maintained at ≤ 5 kΩ. Resting state EEG was recorded for three minutes during eyes-open and three minutes during eyes-closed conditions. During the eyes-open condition, participants were instructed to fixate their eyes at a crosshair located at the center of the monitor.

### EEG analyses

EEG data analysis was conducted using EEGLAB version 2020.0 (Swartz Center for Computational Neuroscience, University of California at San Diego, CA) and MATLAB R2019b (Mathworks Ltd., Natick, MA). Raw EEG data were first filtered using a bandpass filter between 1-50 Hz. We utilized ICA algorithm in EEGLAB to identify and correct for eye blinking, muscle activity, line noise, and motion-related artifacts. Then, EEG data were re-referenced using the Common Average Reference (CAR). The noise-free periods in the EEG recordings were selected by visual inspection and sliced into two-second epochs. Fast Fourier Transform (FFT) was applied to preprocessed EEG to calculate signal power of EEG, i.e., power spectra (µV^2^). Average spectral power in the epochs were calculated for the following frequency bands: Delta (1-4 Hz), Theta (4-8 Hz), Alpha (8-13 Hz), Beta (13-30 Hz), and Gamma (30-50 Hz). From the initial participant pool, 2 smokers, 3 past-smokers, and 5 non-smokers were excluded from further analysis due to excessive EEG artifacts (> 75 μV throughout the entire EEG or EEG spectral power deviated > 3 SD from the group mean in each condition). In the remaining participants, EEG data with the spectral power deviated > 3 SD from the group mean in a single condition (i.e., eyes-open or eyes-closed) were only excluded from the statistical analysis for the corresponding condition: 5 smokers, 6 past-smokers, and 3 non-smokers.

To examine impacts of smoking status among groups, we calculated relative spectral powers, alpha reactivity, and alpha band coherence. The relative spectral power of each frequency band was defined as a ratio between the absolute power of the corresponding frequency band and the total sum of absolute power from 1-50 Hz. To examine alpha band reactivity, we calculated EEG band power changes between eyes-closed and eyes-open conditions as follows [32]:

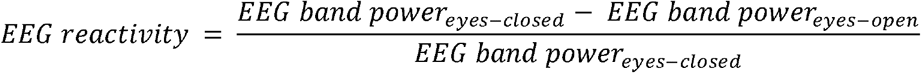

According to this definition, individuals who show larger alpha power difference between eyes-open and closed conditions have larger EEG reactivity.

The alpha band coherence was defined as the magnitude-squared coherence of the two signals using Welch’s mean-corrected periodogram [44]. Given the two stationary signals *x* and *y*, the magnitude-squared coherence is defined as follows:

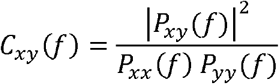

where *P*_*xx*_*(f)* and *P*_*yy*_*(f)* are the power spectral density of signals *x* and *y*, respectively, and *P*_*xy*_*(f)* is the cross power spectral density of the two signals. The alpha band coherence was calculated between all pairs of EEG channels in alpha frequency band.

### Statistical analyses

We conducted one-way analysis of variance (ANOVA) with post-hoc Tukey’s tests for comparing age, income levels, and self-report depression levels (K-BDI-II) among the groups, and used the chi-squared test for comparing education levels among the groups. To test whether smokers and past-smokers differed on their smoking behaviors and self-report attitude against nicotine (dependence and craving), we used independent sample t-tests and compared age of smoking initiation, years of smoking, average number of cigarettes smoked per day, nicotine dependence (FTND total score), and nicotine craving (QSU-brief score). Mann-Whitney U test was used in comparing the number of quit attempts between smokers and past-smokers, because the data violated assumptions of normality. Group differences in EEG band powers, alpha reactivity, and alpha band coherence were evaluated using Kruskal–Wallis test, a non-parametric test equivalent to one-way ANOVA. In all tests comparing EEG features, significance level was set at p-value < 0.05 after controlling for False Discovery Rate (FDR) across 31 channels, and post-hoc Tukey’s tests were conducted for comparison between groups. To further examine the associations between EEG features and individuals’ smoking-related characteristics, we first averaged each EEG feature across the entire brain as an individual’s representative neural measure (unless stated otherwise), and then evaluated Pearson’s correlations between EEG features and individuals’ self-reported measures related to smoking (e.g., FTND, QSU-brief, number of cigarettes smoked per day). Nonparametric bootstrapping method was used to compute confidence intervals (number of the bootstrapped samples = 10,000).

## Results

### Smokers, past-smokers, and non-smokers were matched for all demographics but smoking-related measures

Participants across three groups were matched for age (F(2, 78) = 1.76, *P* = 0.18), gender (all male), education (χ^2^(8) = 14.37, *P* = 0.073), and income level (F(2, 78) = 0.37, *P* = 0.69; **Table 1**). Smokers and past-smokers differed in their smoking-related characteristics. Compared with past-smokers, smokers initiated smoking at their earlier age (t(46) = -2.95, *P* = 0.0049), smoked for more years (t(46) = 13.63, *P* = 9.07e-18), reported higher nicotine dependence (FTND: t(37) = 2.81, *P* = 0.0079), and higher craving at the time of experiment (QSU-brief: t(37) = 4.46, *P* = 7.30e-05). However, average number of cigarettes that individuals smoked (or used to smoke) per day (t(39) = 0.47, *P* = 0.64) as well as the number of quit attempts (U = 142, *P* = 0.12) were comparable between the two groups. Besides these basic demographic information and smoking-related measures, we also collected individuals’ self-report depression symptoms that may accompany their smoking history, and confirmed that all three groups showed comparable levels of depression (F(2, 78) = 0.35, *P* = 0.70). These results indicate that any group differences, if exist, can be attributed to differences in smoking status among groups rather than other comorbidities.

### Low frequency band powers and alpha reactivity are associated with individuals’ smoking status and habits

To test whether EEG powers are associated with individuals’ chronic smoking and abstinence, relative spectral powers were calculated in each frequency band for each group of individuals. In the eyes-open condition, there was no significant group difference in EEG relative band powers. On the contrary, for the eyes-closed condition, significant group differences were found in theta and alpha bands (Kruskal-Wallis test, false discovery rate adjusted *P (P*_*FDR*_) < 0.05; **Fig. 1**). Specifically, relative theta band power was significantly lower in smokers compared with non-smokers in a widespread area including the channels from the frontal, central, parietal, and occipital regions (Fp1, F3, Fz, FT9, FC1, FC2, FC5, C3, C4, TP9, CP1, CP2, CP5, CP6, P3, P4, P7, P8, Pz, O1, O2, and Oz; *P* < 0.05, post-hoc Tukey’s test). Past-smokers also showed diminished relative theta band power, such that their theta band powers from the frontal and occipital regions (Fp1, F4, F7, F8, FT9, FC6, P4, O2, and Oz; *P* < 0.05 in post-hoc Tukey’s test) were significantly lower than that of non-smokers. No significant theta power differences were observed between smokers and past-smokers.

**Figure 1.**
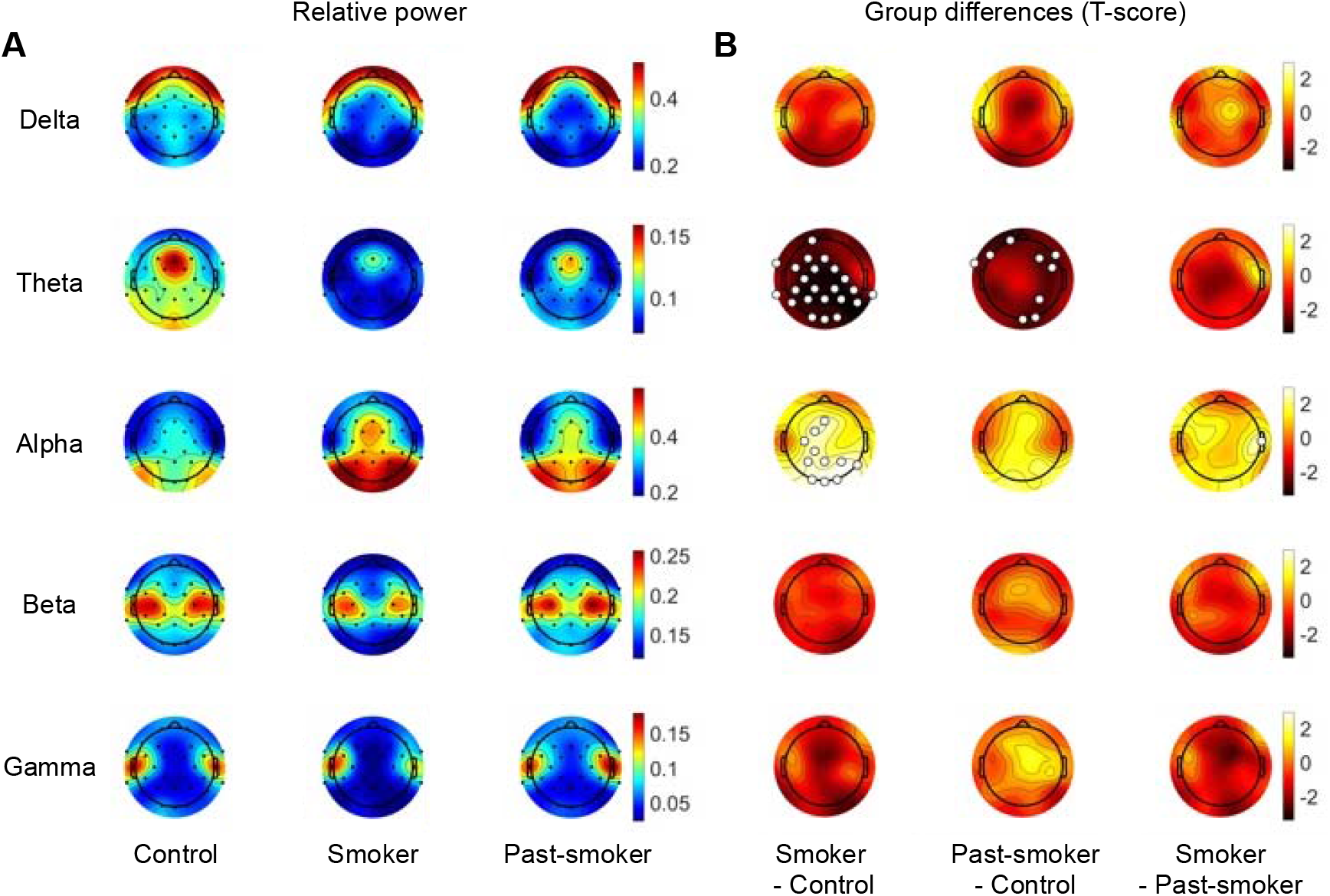
Relative EEG band power (eyes-closed condition). **(A)** Each row illustrates the average relative band powers calculated in each group. **(B)** Statistical group differences in relative band powers were observed in theta and alpha bands, particularly between the Smoker and the Control groups. Specifically, non-smoker controls showed larger relative power in theta band compared to the smoker and past-smoker groups, while smaller power in alpha band than the smoker group. The channels that showed a significant group difference are marked with larger unfilled markers (*P*_FDR_ < 0.05).

In relative alpha band power, significant group differences were largely observed between the smoker and non-smoker groups. Smokers exhibited higher alpha power compared with non-smokers in the central and occipital regions (Fz, FC1, C3, CP1, P3, P4, P8, Pz, O1, O2, and Oz; *P* < 0.05, post-hoc Tukey’s test). Alpha band power in past-smokers was comparable with that of both the other two groups in all regions, except in T8 channel where smokers showed higher power than past-smokers (*P* < 0.05, post-hoc Tukey’s test).

It is worth noting that group differences in low frequency powers associated with smoking status were only observed from the eyes-closed condition, and that such a discrepancy mirrors previous reports regarding alpha reactivity—power reduction at eyes-open compared to eyes-closed condition within a specific frequency band [31-33]. To directly test our hypothesis about its relevance to chronic smoking, we examined group differences in alpha reactivity. As hypothesized, smokers’ alpha reactivity was significantly higher than that of non-smokers and past-smokers (Kruskal-Wallis test: *P*_*FDR*_ < 0.05; **Fig. 2**). Post-hoc Tukey’s test revealed that smokers showed larger alpha reactivity than non-smokers in a broad area (FP1, F3, F4, F7, Fz, FC1, FC2, FC5, FC6, C3, C4, CP1, CP2, CP6, TP1, P3, P4, P8, Pz, O1, O2, and Oz; *P* < 0.05, post-hoc Tukey’s test). Moreover, a part of the increased alpha reactivity in smokers, observed from the central and parietal regions (FC5, FC1, P8, CP6, CP2, and C4; *P* < 0.05, post-hoc Tukey’s test), was significantly higher than that in past-smokers. For completeness, we also calculated EEG reactivity in other frequency bands. However, there was no significant reactivity difference in other frequency bands between groups.

**Figure 2.**
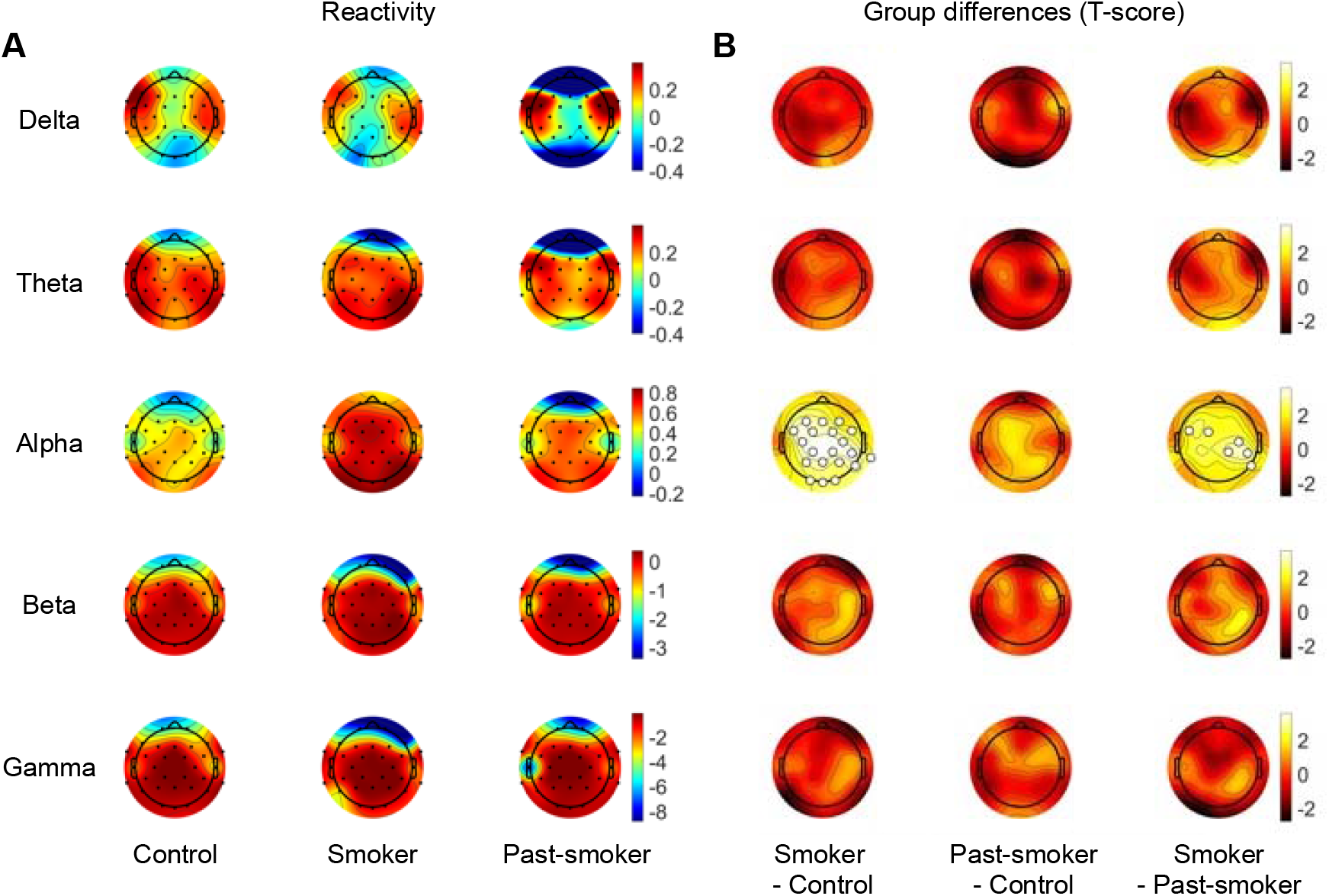
EEG reactivity between the eyes-closed and the eyes-open conditions. **(A)** Each row illustrates average EEG reactivity measures calculated in each group, for each frequency band. **(B)** Statistical group differences in EEG reactivity were observed in alpha band. Particularly, smokers showed larger alpha reactivity compared with past-smokers or non-smoking controls. The channels that showed a significant group difference are marked with larger unfilled markers (*P*_FDR_ < 0.05).

We further examined whether these features in low frequency bands are associated with individuals’ smoking-related characteristics other than their smoking status. Across the two smoker groups (smokers and past-smokers), the amount of cigarette consumption, nicotine dependence, and the number of quit attempts were significantly correlated with the relative powers in the theta frequency band (**Fig. 4**). Particularly, individuals who smoked more number of cigarettes per day (Pearson’s correlation coefficient r = -0.28, bootstrapped *P* = 0.039) or reported higher nicotine dependence (FTND: r = -0.40, bootstrapped *P* = 0.002) showed lower theta band power, while individuals who had more attempts to quit smoking showed higher theta band power (r = 0.43, bootstrapped *P* = 0.033). A significant association with individuals’ nicotine dependence was also observed in the alpha band power, averaged over the occipital regions (O1, Oz, and O2). Individuals who reported higher nicotine dependence showed higher alpha band power (r = 0.30, bootstrapped *P* = 0.031). Although statistically not significant, other two smoking-related measures (cigarettes per day, quit attempts) showed trending associations consistent with the pattern observed in theta band power, considering the opposite relationship theta and alpha powers showed among the groups; individuals who smoked more cigarettes or had less quit attempts tend to show higher alpha power (cigarettes per day: r = 0.16, bootstrapped *P* = 0.30; quit attempts: r = -0.12, bootstrapped *P* = 0.46). No significant correlation was found between alpha reactivity and smoking-related self-report measures. These results indicate that low frequency band powers measured from eyes-closed rsEEG not only project individuals’ smoking status, but also capture detailed individual differences in their nicotine dependence severity.

### Alpha coherence is associated with individuals’ smoking status

To test whether EEG coherence is associated with smoking as observed in other substance use disorders, we examined group differences in alpha band coherence across every pair of channels. As in the aforementioned patterns of EEG powers, significant group differences were observed from the coherence during the eyes-closed condition (Kruskal-Wallis test: *P*_*FDR*_ < 0.05; **Fig. 3**). Specifically, smokers had significantly higher alpha band coherence compared to non-smokers, primarily at the channel pairs among the left frontal, left parietal, and occipital regions (**Fig. 3B, C**). Among the alpha band coherence in smokers, coherence between the left and right frontal regions was higher than corresponding coherence patterns in past-smokers (**Fig. 3B, C**). Coherence in past-smokers was largely comparable with that in non-smokers, except for a few pairs of channels between the left frontal and occipital regions (**Fig. 3B, C**). For completeness, we also examined EEG reactivity in other frequency bands, albeit no significant group difference was observed.

**Figure 3.**
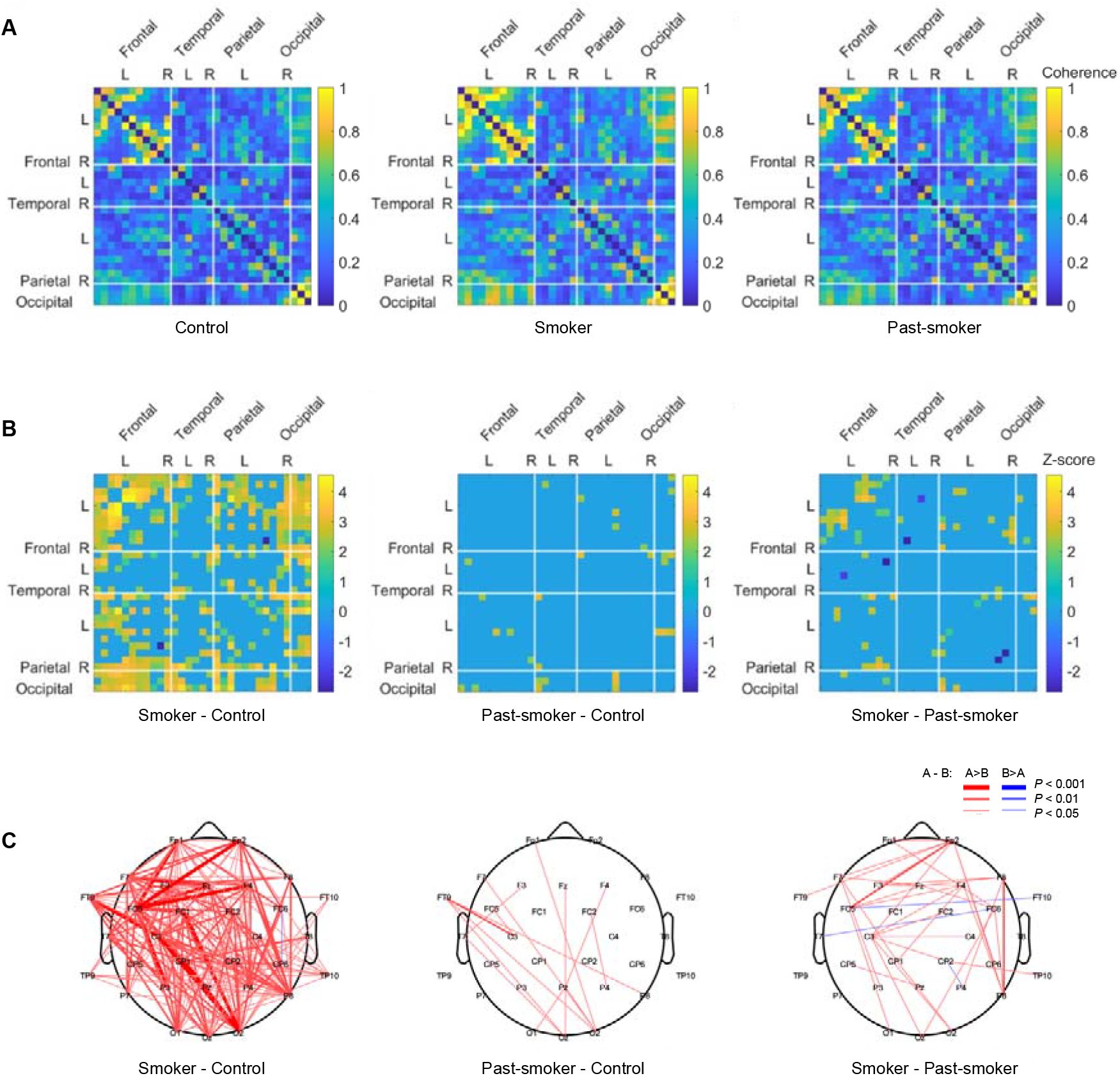
Group differences in alpha band coherence. **(A)** Alpha band coherence between EEG channels was calculated in each group. **(B)** Significant group differences in alpha band coherence were observed most distinctively between the Smoker and the Control groups. **(C)** For an illustrative purpose, all significant group differences in inter-channel coherences are depicted with three different thickness levels (*P*_FDR_ < 0.05, 0.01, and 0.001).

We further examined correlations between individuals’ coherences and their smoking-related measures to test whether the increased coherences are impacts of individuals’ smoking history and/or severity. Regardless of smoking status, individuals who smoked more showed significantly higher alpha coherence (r = 0.40, bootstrapped *P* = 0.026; **Fig. 4C**), evidencing for the severity effect on individuals’ EEG. On the contrary, there was no significant evidence suggesting the impacts of individuals’ self-report nicotine dependence (r = 0.24, bootstrapped *P* = 0.23), quit attempts (r = -0.12, bootstrapped *P* = 0.56), or chronicity (r = 0.31, bootstrapped *P* = 0.12). Consistent with previous findings in individuals with problematic substance uses (e.g., alcohol) [28], these coherence results suggest that chronic and intensive smoking may alter the brain’s functional connectivity, which in turn increases coherences in the rsEEG.

**Figure 4.**
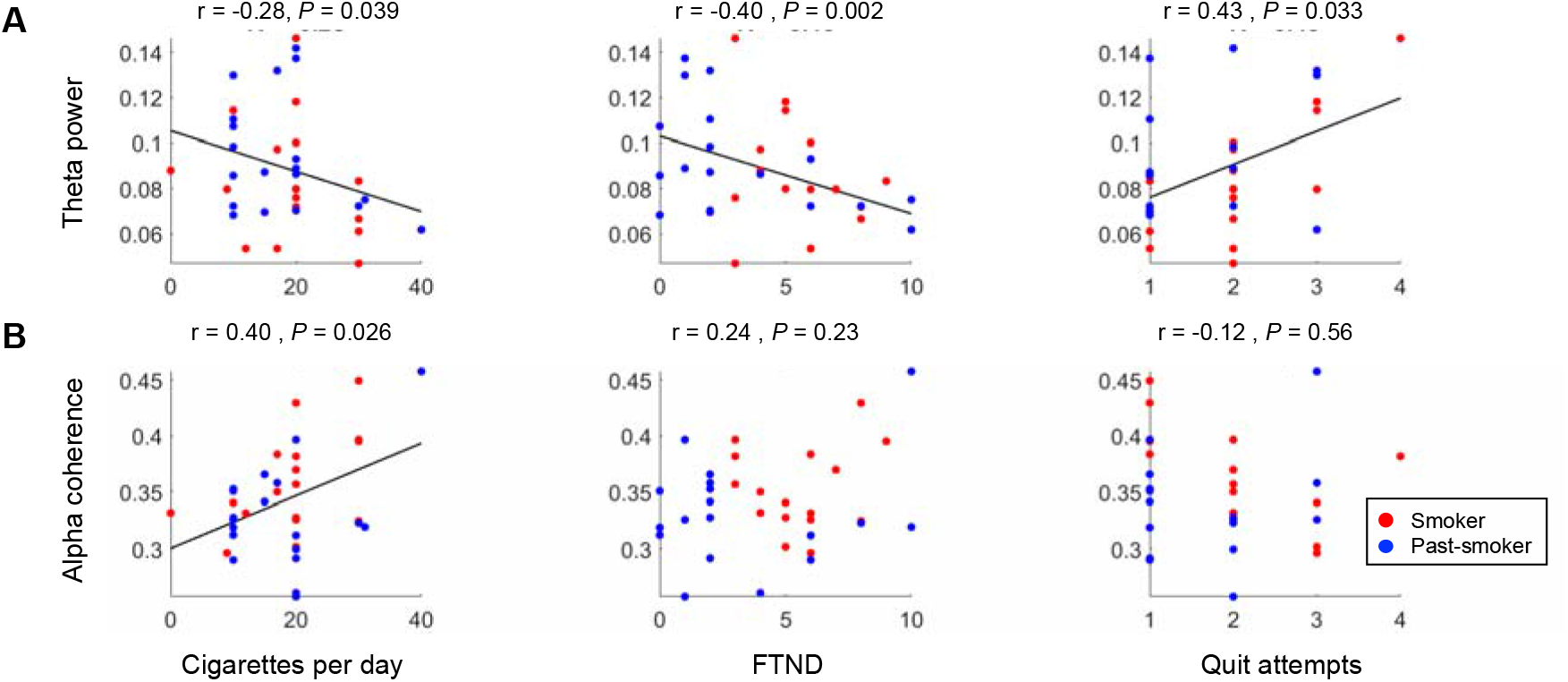
Correlations between EEG features and individuals’ smoking-related characteristics. Pearson’s correlation coefficients were calculated to examine the association between EEG characteristics (relative EEG band powers and coherence) and individuals’ smoking history measures (cigarettes per day, FTND, and the number of quit attempts). **(A)** Across smokers and past-smokers, individuals who have smoked more (or had smoked more in the past for past-smokers) showed lower theta band power, **(B)** but higher alpha coherence. **(A)** In addition, individuals who self-reported higher nicotine dependence (FTND score) showed lower theta band power, and who had more quit attempts showed higher theta power. Each dot represents an individual participant, and the color-coded lines are the regression line between the corresponding measures.

## Discussion

The current study examined three groups of individuals with different smoking profiles and investigated both the long-lasting and reversible effects of smoking on the brain. We compared resting state EEGs between the two smoker groups and non-smokers to characterize long-lasting effects of smoking, and found that individuals who ever smoked showed lower theta band power. Suggesting for reversible effects of smoking on the brain, current-smokers showed higher alpha band power, higher alpha reactivity, and higher alpha band coherence compared with non-smoking controls, but such differences were not observed between past- and non-smokers. In addition, individuals’ self-report smoking behaviors and dependence accounted for the individual differences in the band powers and coherence across current-and past-smokers. These results delineate smoking induced chronic functional changes apart from transient changes in the brain.

Supporting the widely accepted view of addiction as a brain disease [5,45], long-term exposure to addictive substances, regardless of their legality, is known to trigger functional and structural changes in the brain [46]. Based on these neurobiological changes, pharmacological targeting [47] and brain stimulation [48] have been suggested as plausible treatments for reversing the impacts of addiction on the brain. To date, however, previous studies focused on either the impacts of brief abstinence or that of acute nicotine consumption [49], and it still has not been directly examined whether the neural hints of individuals’ smoking history in the brain *ever* disappear even after a long period of successful abstinence. The current study addresses this gap by including past-smokers who smoked for at least six months and stayed abstinent for longer than 20 years. Moreover, smokers were recruited from a group behavior therapy for smoking cessation, and all the participants in the smoker group were mandated to be free from the recent influence of nicotine at the time of experiment. These settings matched sobriety of the two smoking groups. Thus, the differences we report cannot be interpreted as results of instant satiety in the smoker group, but rather as patterns distinguishing the long-lasting impacts of smoking from the impacts of chronic smoking.

Previous studies reported that nicotine-deprived smokers compared to non-smokers or to non-deprived smokers showed reduced power in low frequency bands (i.e., delta and theta) [16,25]. In contrast to these results, another study that specifically examined rsEEG of non-deprived smokers observed a similar pattern of reduced low frequency power compared to that of non-daily smokers [50]. This finding suggested that the changes in rsEEG might not simply be reflecting a state of withdrawal, but also associated with categorical smoking status (e.g., daily smoker, nondaily smoker). As noted above, all participants in the current study were abstinent, and thus any observed differences could be attributed to individuals’ smoking history (e.g., severity, quit attempts, dependence) and status (e.g., current-, past-, and non-smoker). The group difference between current- and past-smokers, which we reported as long-lasting impacts of smoking, was consistent with previous results, such that theta band power in the frontal and occipital regions was lower for current-smokers. Beyond this group difference, these patterns were more distinctive in individuals who have (or had) smoked more number of cigarettes per day, had less quit attempts, and had higher nicotine dependence. These results suggest that reduced low frequency power is a neural marker reflecting both individuals’ smoking history and dependency, and that the ‘scar’ remains through a long abstinence.

Another set of features in alpha band indicated the impacts of chronic smoking. Specifically, under eye-closed condition, current-smokers showed increased alpha band power compared with non-smokers. There was no significant group difference between current- and past-smokers, while the association between individuals’ nicotine dependence and their alpha power suggested that past-smokers are at an intermediate state in-between the other two groups. Complementary to these results, smokers showed similar patterns in their alpha reactivity measure, such that current-smokers showed the most distinctive and highest alpha reactivity, and past-smokers showed an intermediate level of alpha reactivity. Unlike previous studies that examined the effects of acute administration of nicotine [24,25,36], these results provide converging evidence for neural changes accompanied by chronic smoking. Long-term nicotine exposure induces upregulation of nicotinic acetylcholine receptors and their desensitization [35,51,52], which may underlie the changes in functional connectivity between cortical regions and the NBM [32], and in turn cortical activities and EEG changes (e.g., alpha reactivity) [53]. On the contrary to a previous study that reported normalization of nAChR availability from smokers who stayed abstinent for 6-12 weeks [54], our results from past-smokers who successfully abstained for more than 20 years suggest more persistent effects of smoking. Future study may directly collect both EEG and metabolic imaging (e.g., single-photon emission computed tomography; SPECT) to confirm these long-lasting effects.

Relatively little is known about the impacts of smoking on EEG coherence [49]. One previous study, while the sample size was small, reported a consistent pattern in alpha coherence, such that smokers showed higher coherence than age-matched non-smokers [55]. Our data replicate and expand this result. Current-smokers showed higher alpha coherence than non-smokers across the whole brain, and moreover, past-smokers still showed higher coherence compared with non-smokers between the frontal and occipital regions. Most interestingly, such an alteration in alpha coherence was associated with the number of cigarettes individuals smoked a day, regardless of their smoking status; individuals who smoked the most cigarettes a day, even after 20 years of abstinence, showed the highest alpha coherence. These results support that not only can EEG coherence be used as a biomarker detecting substance use disorder (e.g., cannabis use [27], internet addiction [30]), but also as a measure reflecting a trace of long-term substance uses (e.g., alcohol use [28]).

The current study has the following limitations. First, it is a cross-sectional study and thus, we cannot answer whether the observed neural patterns (e.g., higher alpha power) are indeed consequences of chronic substance use (or abstinence) or individuals’ traits that should get credits for making them more (or less) prone to be addicted to substances. Future study that uses longitudinal tracking of individuals’ substance use history in conjunction with their neural data may provide further insights about causal roles of their brain’s characteristics in initiation of substance dependence [56] as well as in sustained remission [57]. Second, the current study only includes data from male participants, and thus leaves it untested whether the same EEG features can be used in characterizing female smokers. Previous studies suggested that female smokers have different smoking behavior [58] and moreover, different nicotine metabolism [59]. Given these reports, it is reasonable to expect that female smokers may show different neural signatures along their chronic smoking and abstinence. Direct investigation about and comparison between smokers of all genders would further expand our understanding of why there is a higher percentage of male smokers in general [60] and why some individuals succeed better at quitting.

To conclude, we investigated how chronic smoking and long-term sustained remission affect the brain. Our data show that the features observed from chronic smokers’ resting-state EEG remain at a significant level even after more than 20 years of sustained remission, which suggests long-lasting and potentially irreversible impacts of smoking. It has been already well known that smokers who attempt to quit are highly likely to relapse and there are various factors associated with their failures [2,3]. Most of these risk factors (e.g., nicotine dependence, social environment) predicted relapse within the first few months of quitting, but failed to predict why some individuals still relapse after a few years of abstinence [61]. Our findings provide a neural explanation why individuals who ever initiated smoking in a lifetime may have changes in their brain and show different neural responses to perceptual [11] and social information [62], and also might be at constant risk of further problem behavior [63].

## Data Availability

All data produced in the present study are available upon reasonable request to the corresponding authors.

## Acknowledgements

This work was supported in part by UNIST internal funding (1.210046.01) and the National Research Foundation of Korea (NRF-2018R1D1A1B07043582 to Chung). K.B. was partly supported by the National Research Foundation of Korea (NRF-2021R1F1A1063968), Institute of Information & Communications Technology Planning & Evaluation (grant No. 2020-0-01450, Artificial Intelligence Convergence Research Center [Pusan National University]), and Ministry of Education of South Korea (the BK21 Four program, Korean Southeast Center for the 4th Industrial Revolution Leader Education).

## Author contributions

Conceptualization: D.C.; Methodology: H.L., Y.J., C.Y., K.B., and D.C.; Formal analysis: H.L., Y.J., and K.B.; Funding acquisition: K.B. and D.C.; Investigation: H.L., Y.J., H.S., J.P., M.H., K.B., and D.C.; Writing – original draft: H.L., Y.J., K.B., and D.C.; Writing – review and editing: H.L., Y.J., C.Y., H.S., J.P., M.H., K.B., and D.C.; Supervision: K.B. and D.C.

## Competing interests

The authors declare no competing interests.

## Notes

### Competing Interest Statement

The authors have declared no competing interest.

### Author Declarations

The current study and all the protocols were approved by the Institutional Review Board (IRB) of Ulsan National Institute of Science and Technology (UNIST) (UNISTIRB-19-45-A), and all participants provided written informed consent following an explanation of study procedures.

